# Oncological outcomes of primary renal malignancies other than clear cell renal carcinoma: A retrospective study from a tertiary center

**DOI:** 10.1101/2020.11.27.20239889

**Authors:** Ketan Mehra, Manikandan Ramanitharan, Dorairajan Lalgudi Narayanan, Sreerag Kodakkattil Sreenivasan, Sidhartha Kalra

## Abstract

**Introduction:** A lot of research is available about clear cell renal carcinomas (ccRCC). But there are lesser known facts about other subtypes of renal malignancies. With advances in immunohistochemical and cytogenetic techniques, new variants of renal tumors are being increasingly reported. The treatment and prognosis of such rare malignancies is still an enigma. We performed this study to analyze the incidence, clinico-pathological features, surgical treatment, and survival of non-clear cell RCC at our institution.

**Materials and Methods:** The histopathological reports of 77 Nephrectomy specimens who underwent surgical treatment for suspected renal tumors from 2013-2018 were retrospectively reviewed. 19 (24%) of patients had documented uncommon histologic variants of RCC. The clinical, demographic, and histologic characteristics of these patients were analyzed, and survival was evaluated. The characteristic light microscopy and immunohistochemical features of these lesions were documented.

**Results:** Mean age was 45 years (21-67 years). Out of 19 patients, 14 (73.6%) were males, and 5 (26.4%) were females. Mean tumor size was 12 (6-31) cm in the largest dimension. 17 (22%) patients underwent radical nephrectomy, and 2 (2.5%) were treated with partial nephrectomy. Patients with collecting duct, synovial sarcoma, and primitive neuro-ectodermal tumor (PNET) had associated inferior vena caval thrombus and underwent venous thrombectomy. Adjuvant treatment in the form of chemotherapy was instituted in collecting duct, adult Wilms and pure sarcomas. There was no mortality in the papillary carcinoma, and the worst prognosis was encountered in

**Conclusion:** Sarcomatoid and collecting duct variants were associated with worse prognosis. Presently, aggressive surgical extirpation is the mainstay in the management of these histologic variants. Adjuvant chemotherapy and TKI inhibitors have a limited role.

## Introduction

Renal tumors are a cause of significant morbidity and mortality worldwide. The advances in radiological imaging and its widespread adoption have translated into earlier detection leading to better outcomes. As per WHO classification, renal tumors can be classified as renal cell tumors, metanephric tumors, mixed mesenchymal and epithelial tumors, nephroblastic tumors, neuroendocrine tumors, lymphomas, and metastatic lesions to the kidneys.^[1]^ The incidence of histological classification of renal cell carcinomas (RCC) include Clear cell carcinoma 70%, papillary 10-15%, chromophobe 4-6%, Collecting duct < 1% and unclassified lesion 4-5% (Table 1).^[2]^ Sarcomatoid differentiation occurs in 1-5% of malignant renal tumors and portends a poorer prognosis.^[3]^ Different histologic types have a different biologic and prognostic profile. Among these, clear cell RCC has a less favorable prognosis. Papillary and chromophobe variants have a preferably good prognosis. Still, others like angiosarcoma, collecting duct carcinoma, renal medullary carcinoma, and sarcomatoid variants have a poorer prognosis.^[4, 5]^ The aim of the present study is to analyze the incidence, clinico-pathological features, surgical treatment, and survival of non-clear cell RCC at our institution.

**Table 1.** Incidence of uncommon renal tumors.

| **Type of cancer** | **Literature review** | **Our study (%)** |
| --- | --- | --- |
| Clear Cell carcinoma | 70% | 52 (67.5) |
| Papillary RCC | 10-15%, | 5 (6.5) |
| Chromophobe RCC | 4-6%, | 4 (5.2) |
| Collecting duct RCC | < 1% | 3 (3.9) |
| Primary angiosarcoma<sup>[16]</sup> | < 40 cases | 1 (1.3) |
| Primary synovial sarcoma<sup>[14]</sup> | 34 cases | 1 (1.3) |
| Adult Wilms tumor<sup>[15]</sup> | <1% | 1 (1.3) |
| Unclassified renal sarcoma | - | 1 (1.3) |
| Primitive Neuro-ectodermal (PNET) tumor<sup>[18]</sup> | 80 cases | 1 (1.3) |
| Sarcomatoid differentiation of different renal tumors<sup>[3]</sup> | 1-5 % | 3 (3.9) |

RCC: Renal cell Carcinoma

## Materials and Methods

### Data collection and study design

The study included 77 renal tumor patients who underwent radical or partial nephrectomy from January 2013 to July 2017 in a tertiary care center in South India. The patients’ records were retrospectively reviewed for demographic, radiologic, and pathologic data, surgical intervention, and adjuvant therapy if any. 19(24%) out of 77 patients had histopathology reported as uncommon non-clear cell renal tumors.

All the patients presented to the Department of Urology with the complaints of either abdominal or back pain. Besides, two patients had a history of haematuria, and one had significant weight loss and weakness. One patient presented with retroperitoneal hemorrhage (Wunderlich syndrome) and shock and underwent partial nephrectomy after stabilization. All the patients initially underwent ultrasonography, which diagnosed renal tumors. Fifteen patients underwent a CECT abdomen, and three had an MRI abdomen for accurate staging and surgical planning.

## Results

### Tumor characteristics

Out of 19 patients, 14 (73.6%) were males, and 5 (26.4%) were females. The mean age was 45 (range 21-67) years. Abdominal USG revealed the presence of a heterogeneous mass in the involved kidney in all the patients. On cross-sectional imaging, the mean tumor size was 8.8 (range 6-36) cm in the largest dimension. Three out of 19 had venous thrombosis (Collecting duct carcinoma, synovial sarcoma, and primitive neuro-ectodermal tumor) and underwent MRI to stage tumor thrombosis. Three patients had clinically significant lymph node enlargement. On histopathological evaluation, the following histologies were identified: Papillary Carcinoma-5, chromophobe-3, Collecting duct carcinoma-3, clear cell with sarcomatoid differentiation-2, chromophobe with sarcomatoid differentiation-1, primary renal sarcoma-5 [primary biphasic synovial sarcoma-1, Angiosarcoma-1, unclassified sarcoma-1, Adult Wilms tumor-1, primitive neuro-ectodermal tumor (PNET)-1]. The light microscopy and immunohistochemical features of these lesions were documented (Table 2). Among these, 10(53%) patients had T2 disease, 7 (37%) had T3, and 2 (10%) patients had T4 disease. Two patients had metastatic lymph nodes positive histopathologically (collecting duct carcinoma and Synovial sarcoma), and the same patients had venous tumor thrombus. These findings were also diagnosed radiologically.

**Table 2.** Microscopic features of different subtypes of renal tumors.

| **S.No.** | **Type of carcinoma** | **Type of cells** | **Immunohistochemistry** |
| --- | --- | --- | --- |
| 1 | Papillary RCC | Pseudostratified cells with abundant cytoplasm having atypical nuclei with prominent nucleoli | **CK7** and **CD10** negative |
| 2 | Chromophobe RCC | Finely reticular pale cytoplasm, perinuclear halos, and wrinkled raisinoid nuclei. | **CD117** positive.<br>**CK7** focally positive.<br>**Vimentin** Negative |
| 3 | Collecting duct RCC | Polygonal shaped cells with squamous differentiation and abundant keratin pearls | **CK 5/6, Pancytokeratin,** and **Vimentin** positive.<br>**CD10, CD117** Negative |
| 4 | Chromophobe with Sarcomatoid Variant | Abundant granular eosinophilic cytoplasm with admixed spindle cells. Bizarre cells present. | **CD-117** positive in kidney chromophobic component.<br>**CK-7** focally positive.<br>**Vimentin** and **SMA** positive in spindle cells. |
| 5 | Angiosarcoma | Polygonal/spindle-shaped cells. Moderate vesicular nuclei present eosinophilic cytoplasm. | **CD34, Factor 8, and CD31** positive.<br>**INI-** Negative |
| 6 | Biphasic Synovial Sarcoma | Spindle cells with marked nuclear atypia with mast cell infiltration. Marked Pleomorphic nuclei. | **Cytokeratin, CD99, S100,** and **Bcl-2** positive |
| 7 | Adult Wilms tumor | Highly cellular with blastemal, stromal, and epithelial derivatives. Elongated, wedge-shaped nuclei with frequent mitoses. | **Vimentin** and **PanCk** positive.<br>**EMA** and **Bcl2** focal positive.<br>**CD10, CD99, CD56, synaptophysin, CK7, WT 1, Desmin, and myoD1** negative. |
| 8 | Unclassified Sarcoma | Tumor cells appeared small, round to oval-shaped, with scant cytoplasm vesicular chromatin and tiny nucleoli with extensive hemorrhagic areas with hemosiderin-laden macrophages. | **Vimentin, BCL2** and **FL1** positive.<br>**Pan CK, CK7, CD10, CD31, S100, HMB45, SMA, ER, PR, and WT-1** negative |
| 9 | PNET | Nests of cells intersected by fibrous septa present. Tumor cells show hyperchromatic nuclei, inconspicuous nucleoli, and scant cytoplasm with brisk mitosis and necrosis is present. | **CD-99** positive.<br>**WT 1, CK, desmin, BCL-2, FLI 1** negative. |

CK- Cytokeratin
CD- Cluster of Differentiation
SMA- Smooth muscle antigen

### Treatment

Out of 19, 17(89%) patients underwent radical nephrectomy, and 2(11%) had partial nephrectomy. At the time of relapse, patients with RCC except collecting duct were given targeted therapy in the form of sorafenib. Collecting duct carcinoma, which relapsed, were given chemotherapy Gemcitabine and Cisplatin. Sarcoma patients were given adriamycin, cisplatin, ifosfamide based chemotherapy.

### Survival

Of the 70 patients with malignant tumors, 29 died (41.4%) during the follow-up period; the death rates in males and females were 22.8% and 18.1%, respectively. At a median follow-up of 24 months, 21(40.4%) patients with clear cell subtype died. Papillary cell carcinoma had a good outcome with all the patients alive. Among three patients of Chromophobe renal cell carcinoma, 1(33.3%) patient died. 2 out of 3(66.6%) patients of collecting duct carcinoma died. Sarcomatoid differentiation also had the same mortality rate; 2 out of 3 (66.6%) succumbed to the disease. One patient of PNET died in the perioperative period. Among the four with primary renal cell sarcomas, patients with angiosarcoma and adult Wilms tumor histology died.

## Discussion

On reviewing literature, clear cell carcinoma, which is the most prevalent of all malignant tumors, has poorer survival as compared to papillary and chromophobe variants.^[6]^ Different studies showing the prognosis of uncommon renal malignancies are shown in Table 3. Chromophobe renal cell carcinoma (RCC) tends to occur in the 6^th^ decade of life and has a relatively better prognosis than other subtypes of RCC.^[2]^ Choueiri et al. suggested that sunitinib and sorafenib are effective in metastatic chromophobe RCC as in their study, 75% of patients had stable disease for more than three months, and 25% reported a partial response.^[7]^ According to Sterc et al., chromophobe RCC cells have overexpression of the CD117 marker. Hence, targeted agents like imatinib, dasatinib, and nilotinib may still have a role in their treatment as adjuvant agents.

**Table 3.** Different studies showing survival and prognosis in uncommon renal malignancies.

| **Subtype of Renal tumors in the study** | **Study** | **Outcome** | **Treatment given** |
| --- | --- | --- | --- |
| Metastatic non-clear Cell Carcinoma | Agarwala et al.<sup>[19]</sup><br>(n=40) | OS for :<br>1. Papillary 9.8 months,<br>2. Chromophobe 30.3 ± 8.4 months<br>3. Sarcomatoid 4 months.<br>4. Others for 7.9 months. | Either of them as first-line therapy:<br>1. Sorafenib<br>2. Sunitinib<br>3. Pazopanib<br>4. Everolimus<br>5. Supportive care |
| Chromophobe RCC | Klatte et al.<sup>[20]</sup><br>(n=124) | 1. 5-year DSS 78%<br>2. OS- 6 months | Nephrectomy ± Immunotherapy |
|  | Lee et al.<sup>[21]</sup><br>(n=148) | 1. 5-year RFS 82.7%<br>2. 5-year CSS 88.8% | Nephrectomy |
| Collecting Duct RCC | Kwon et al.<sup>[11]</sup><br>(N=35) | 1. PFS-5.8 months<br>2. OS- 54.4 months | Nephrectomy ± Immunotherapy/<br>Chemotherapy/<br>Targeted therapy |
|  | Ciszewski et al.<sup>[12]</sup><br>(n=10) | 1. RFS- 4.9 months<br>2. Time for distant metastases- 8.1 months | Nephrectomy ± Gemcitabine+Cisplatin/<br>Radiotherapy/<br>Metastatectomy |
| Sarcomatoid variant | Cheville et al.<sup>[22]</sup><br>(n=120) | 1. OS-8 months<br>2. 5 year CSS- 14.5% | Nephrectomy |
|  | Shuch et al.<sup>[9]</sup><br>(n=62) | OS- 4.9 months | Cytoreductive nephrectomy ± interleukin-2/<br>Gemcitabine/<br>Doxorubicin |
|  | Mian et al.<sup>[10]</sup><br>(n=108) | OS- 9 months | Nephrectomy ±<br>Systemic therapy |
| Sarcoma | Moreira et al.<sup>[13]</sup><br>(n=489) | 1. In non-metastatic 5-year CSS-58%<br>2. In metastatic 5-year CSS- 6%, | 1. Surgery only<br>2. Surgery+radiation<br>3. Radiation only<br>4. No treatment |
|  | Wang et al.<sup>[17]</sup><br>(n=41) | 1. 5-year survival- 14.5 %<br>2. OS- 28 months | 1. Surgery<br>2. Chemotherapy<br>(Adriamycin,cisplatin-based)<br>3. Radiotherapy |

OS-Overall survival
DSS-Disease specific survival
RFS- Relapse free survival
CSS- Cancer-specific survival

The sarcomatoid variant has been reported to have a median survival of 4-9 months after being diagnosed.^[8]^ Cytoreductive nephrectomy has been an integral part of treatment for advanced conventional RCC. But in cases of sarcomatoid variants, it has a doubtful benefit in survival, with 60% of patients not able to proceed to the stage of targeted therapy after surgery.^[9, 10]^ Therefore Shuch et al. suggested that upfront tyrosine kinase inhibitor (TKI) therapy would be a better option for these patients and reserving surgery for those patients with good performance status and who demonstrate good clinical response.^[8]^ Collecting duct RCC is a sporadic tumor with a <1% incidence and has a poor survival. In a multicentre study, Kwon et al. studied 35 patients of collecting duct carcinoma and reported median progression-free survival (PFS) of 5.8 months and overall survival (OS) of 54.5 months.^[11]^ In their study, 27 patients underwent nephrectomy, comprised of curative surgery in 17 cases, and 10 had palliative surgery. Three patients received upfront chemotherapy, and 4 received no treatment at all. Overall, 22 patient received different palliative chemotherapy regimen in the form of methotrexate, vinblastine, Adriamycin, and cisplatin (MVAC), interferon (IFN), 5-fluorouracil, interleukin-2 (IL-2), gemcitabine, methotrexate, Adriamycin, and cisplatin (GMAC), gemcitabine and cisplatin (GP), gemcitabine and carboplatin (GC). They reported improved overall survival in patients on palliative chemotherapy (18.4 months) than those without treatment (4.5 months). Ciszewski et al., in their study of 10 patients with collecting duct carcinoma, reported a median overall survival of 7.6 months after nephrectomy and chemotherapy or radiotherapy. Only two patients survived > 2 years after the nephrectomy.^[12]^

The primary renal sarcoma is a rare malignancy of kidney, constituting <1% of all malignant renal tumors.^[13]^ Leiomyosarcoma is the most common histological type, followed by liposarcoma, spindle cell sarcoma, malignant fibrous histiocytoma, and fibrosarcoma. Other histological types are rarer like primary synovial sarcoma, adult Wilm’s tumor, and angiosarcoma.^[14-16]^ Due to fewer numbers of cases reported in the literature, the natural history and prognosis of such tumors are difficult to understand. Moreira et al. studied retrospectively 322 non-metastatic patients with primary renal sarcoma and evaluated the survival after surgical resection alone or in combination with radiation.^[13]^ With these modalities, five-year cancer-specific survival was 58%. They attributed age, race, tumor size, and tumor grade to be independently associated with cancer death in non-metastatic disease, while race and tumor histology had prognostic significance in only metastatic disease. Wang et al. reported a high mortality rate of 62.2% at 24 months of follow-up in patients with primary renal sarcoma. Only 8% of patients were disease-free, suggesting the poor prognosis of the disease.^[17]^ They reported a 1, 3, and 5-year survival rate as 86.3%, 40.7%, and 14.5%, respectively.

In our study, we included rare subtypes of adenocarcinoma as well as primary renal sarcoma. Among adenocarcinoma, papillary tumors had a good prognosis with all patients alive at the end of the study period without relapse. Sarcomatoid variants and collecting duct carcinoma had a dismal prognosis, with patients dying within one year of surgery. Four patients of primary renal sarcoma had a varying course. Three patients received adjuvant chemotherapy, out of which two died within two years following surgery. One patient of primary renal sarcoma was managed by partial nephrectomy and is disease-free after 12 months of follow up.

### Limitations of the study

The present study has several limitations. First, the sample size is small, and it is challenging to generalize disease outcomes and prognosis due to less number of cases. Second, it was a retrospective, descriptive study. Well-designed prospective studies with large numbers are needed to study the disease and its prognosis.

## Conclusion

Different subtypes of renal malignancies give prognostic information. At present, aggressive surgical extirpation is the mainstay in the management of histologic variants of RCC. Adjuvant chemotherapy and TKI inhibitors have a limited role. All such variants have lower incidences making it challenging to study the role of adjuvant therapy. Moreover, multi-centric prospective trials with a large patient population are required to assess the impact of newer adjuvant therapies for such rare variants.

## Supporting information

Title page

## Data Availability

The data is confidential and cannot be made available

