## Supplementary material for "Oncological outcomes of primary renal malignancies other than clear cell renal carcinoma: A retrospective study from a tertiary center": Title page

**Type of article:** Original article

**Running title:** Uncommon primary renal malignancies

**Source of Work**: Jawaharlal Intitute of Postgraduate Medical Education and Resarch (JIPMER), Puducherry, India

**Authors:**

1) Dr Ketan Mehra (MS, General Surgery)

Senior Resident, Urology,

Jawaharlal Intitute of Postgraduate Medical Education and Resarch (JIPMER)

Puducherry, India

2) Dr Manikandan Ramanitharan

Additional Professor, Urology,

Jawaharlal Intitute of Postgraduate Medical Education and Resarch (JIPMER)

Puducherry, India

3) Dr Dorairajan Lalgudi Narayanan

Professor, Urology,

Jawaharlal Intitute of Postgraduate Medical Education and Resarch (JIPMER)

Puducherry, India

4) Dr Sreerag Kodakkattil Sreenivasan (MCh, Urology)

Associate Professor, Urology,

Jawaharlal Intitute of Postgraduate Medical Education and Resarch (JIPMER)

Puducherry, India

5) Dr Sidhartha Kalra (MCh, Urology)

Assitant Professor, Urology,

Jawaharlal Intitute of Postgraduate Medical Education and Resarch (JIPMER)

Puducherry, India

**Corresponding author:**

Dr Manikandan Ramanitharan

Additional Professor, Urology,

Jawaharlal Intitute of Postgraduate Medical Education and Resarch (JIPMER)

Puducherry, India

Contact: 9786306473

**Abstract**

**Introduction**

A lot of research is available about clear cell renal carcinomas (ccRCC). But there are lesser known facts about other subtypes of renal malignancies. With advances in immunohistochemical and cytogenetic techniques, new variants of renal tumors are being increasingly reported. The treatment and prognosis of such rare malignancies is still an enigma. We performed this study to analyze the incidence, clinico- pathological features, surgical treatment, and survival of non-clear cell RCC at our institution.

**Materials and Methods:**

The histopathological reports of 77 Nephrectomy specimens who underwent surgical treatment for suspected renal tumors from 2013-2018 were retrospectively reviewed. 19 (24%) of patients had documented uncommon histologic variants of RCC. The clinical, demographic, and histologic characteristics of these patients were analyzed, and survival was evaluated. The characteristic light microscopy and immunohistochemical features of these lesions were documented.

**Results:**

Mean age was 45 years (21-67 years). Out of 19 patients, 14 (73.6%) were males, and 5 (26.4%) were females. Mean tumor size was 12 (6-31) cm in the largest dimension. 17 (22%) patients underwent radical nephrectomy, and 2 (2.5%) were treated with partial nephrectomy. Patients with collecting duct, synovial sarcoma, and primitive neuro-ectodermal tumor (PNET) had associated inferior vena caval thrombus and underwent venous thrombectomy. Adjuvant treatment in the form of chemotherapy was instituted in collecting duct, adult Wilms and pure sarcomas. There was no mortality in the papillary carcinoma, and the worst prognosis was encountered in

**Conclusion:**

Sarcomatoid and collecting duct variants were associated with worse prognosis. Presently, aggressive surgical extirpation is the mainstay in the management of these histologic variants. Adjuvant chemotherapy and TKI inhibitors have a limited role.

**Keywords:** Renal Cell Carcinoma; Nephrectomy; Renal Tumors; Uncommon renal tumors; Renal sarcoma; Collecting duct carcinoma
